# The impact of the Covid-19 lockdown on the experiences and feeding practices of new mothers in the UK: Preliminary data from the COVID-19 New Mum Study

**DOI:** 10.1101/2020.06.17.20133868

**Authors:** Adriana Vazquez, Sarah Dib, Emeline Rougeaux, Jonathan Wells, Mary Fewtrell

## Abstract

**Background:** The COVID-19 New Mum Study is recording maternal experiences and infant feeding during the period of UK lockdown. This report from week 1 of the survey aims to (1) provide information relevant for those supporting this population; (2) identify groups currently under-represented in the survey.

**Methods:** Women living in the UK aged ≥18 years with an infant ≤12 months of age completed an anonymous online survey (https://is.gd/covid19newmumstudy). Information/links are shared via websites, social media and existing contacts.

**Results:** Between May 27^th^ and June 3^rd^ 2020, 1365 women provided data (94% white, 95% married/with partner, 66% degree/higher qualification, 86% living in house; 1049 (77%) delivered before lockdown (BL) and 316 (23%) during lockdown (DL). Delivery mode, skin-to-skin contact and breastfeeding initiation did not differ between groups. DL women had shorter hospital stays (p<0.001) and 39% reported changes to their birth plan. Reflecting younger infant age, 59% of DL infants were exclusively breast-fed or mixed fed versus 39% of BL (p<0.05). Thirteen % reported a change in feeding; often related to lack of breastfeeding support, especially with practical problems. Important sources of feeding support were the partner (60%), health professional (50%) and online groups (47%). 45% of DL women reported insufficient support with feeding. Among BL women, 57% and 69% reported decreased feeding support and childcare, respectively. 40% BL/45% DL women reported insufficient support with their own health, 8%/9% contacted a mental health professional and 11% reported their mental health was affected. 9% highlighted lack of contact and support from family and distress that they had missed seeing the baby.

**Conclusion:** Lockdown has had an impact on maternal experiences, resulting in distress for many women. Survey participants are currently not representative of the population; notably, groups at greater risk are under-represented. Increasing the diversity of participants is a priority.

**Survey funding:** None. All research at Great Ormond Street Hospital NHS Foundation Trust and UCL Great Ormond Street Institute of Child Health is made possible by the NIHR Great Ormond Street Hospital Biomedical Research Centre. The views expressed are those of the author(s) and not necessarily those of the NHS, the NIHR or the Department of Health.

## Introduction

The lockdown measures introduced in the UK on 23rd March 2020 to reduce the spread of Covid-19 resulted in a rapid change in circumstances for mothers and their newborn infants, with the potential for both positive and negative effects. The COVID-19 New Mum Study aims to capture information on maternal experiences and mood, infant feeding practices and perceived infant behaviour during the unique circumstances of the initial lockdown and subsequent stages as restrictions are relaxed. Recording what is happening and understanding the experiences of mothers is key to providing appropriate support. This could prove especially important if a second wave occurs and lockdown restrictions are strengthened again. Considering the impact of lockdown experiences on infant feeding and behaviour from the perspective of maternal-infant conflict, in particular mediated by maternal mood, will also enhance our broader understanding of human lactation^1^.

In this preliminary report, we present descriptive data on the delivery experiences and infant feeding practices of mothers who completed the survey during week 1 (May 27^th^-June 3^rd^, 2020). The aims of this analysis are (1) to provide information that may be of immediate use to health providers and groups supporting pregnant women and new mothers; and (2) to assess whether the sample is representative and identify under-represented groups who should be targeted in the ongoing recruitment strategy.

## Methods

Women currently living in the UK aged ≥18 years who have an infant currently under 12 months of age are being invited to complete a one-time online survey (The Covid-19 New Mum Survey – https://is.gd/covid19newmumstudy). The survey uses the professional software REDCap and can be accessed via a secure webpage from any computer or smartphone with internet access. It takes approximately 15-20 minutes to complete. No personal identifiers are collected, so the data generated are anonymous. Since the duration of the current lockdown and details of the mechanism(s) of exit are currently unclear, the survey will remain open until 31^st^ December 2020. This will allow data to be collected and reported relating to different levels of lockdown restrictions.

Information and links to the survey are being spread via websites and social media groups used by mothers, such as Facebook infant feeding and mother and baby support groups,, Twitter, and Instagram; via existing contacts with relevant professional and support organisations and groups; and via word of mouth.

### Content of the survey

The survey was designed by the study team who, collectively, have experience of research in the field of infant nutrition with backgrounds in paediatrics, child nutrition & dietetics and anthropology. Comments from peer-reviewers were incorporated in the final questionnaire. Care was taken to minimise or avoid questions that might provoke or worsen psychological distress. Where possible, the survey uses questions and formats from previous or ongoing research to facilitate comparisons. Participants have the option to omit questions if they do not want to provide the information requested.

The survey has four parts:

1. *Background characteristics:* Questions address social and demographic factors recognised to impact infant feeding practices, including maternal age and ethnicity, maternal education and household income, household/family composition; and the first three letters of the post-code to give a broad marker of geographical location.
2. *COVID-19 and its impact on household health, work patterns and finances:* Questions ask about experience of COVID-19 and how COVID-19 and the lockdown have affected the household and family, as a measure of the magnitude of the change in maternal circumstances and experience.
3. *Birth experience, infant feeding and behaviour, and changes due to COVID-19*: Questions in this section ask about the birth, infant feeding practices, and sources of information and support. If the baby was born before lockdown (BL) started, the mother is asked how this has changed any aspect of infant feeding, the infant’s behaviour (irritability, sleep, appetite), and access to infant feeding support and childcare support including contact with health care professionals. If the baby was born during the lockdown (DL), the mother is asked if this led her to change her birth and/or feeding plans, and the current support she is receiving.
4. *Impact of COVID-19 on the mother’s activities, mood, concerns and access to support:* Questions ask about how the lockdown has affected maternal activities and mood, considering both positive and negative aspects. This includes the frequency of different activities, such as leaving the home for work, exercise, shopping; engaging in online social activities, or accessing infant support groups. The final section contains questions about the mother’s mood, asking about a range of both positive and negative experiences; and whether she is concerned over her own health, that of her family or friends.

A range of resources and sources of support for both infant feeding and maternal mental health concerns are provided at the end of the survey. The aim is to obtain as many survey responses as possible during the period of lockdown and its subsequent easing, in order to obtain a representative sample and maximise the generalisability and usefulness of the results.

### Ethics and consent

Ethical approval was obtained from the UCL Research Ethics committee (0326/017). The first page of the survey provides information about the study and, having read this, participants are asked to provide consent to participate before proceeding. They are reminded not to provide any information in the free-text boxes that could allow identification.

### Statistical analyses

Data from the survey were exported from REDCap and analysed in SPSS (IBM SPSS statistics v 25). Descriptive data are shown as mean (SD) or n (%). Data are presented separately for women who delivered before (BL) or during the lockdown (DL), with comparisons between groups using Student’s t-test, Mann-Whitney or chi-squared test as appropriate.

## Results

Between May 27^th^ and June 3^rd^, 1457 participants provided complete or partially complete answers to the survey. 1365 women provided data on their delivery experience and infant feeding practices and are included in this analysis. 1049 (77%) gave birth before lockdown and 316 (23%) during lockdown. Characteristics of the participants are shown in Table 1, and their geographical distribution in Figure 1. Notably, 94% of participants self-identified as white, 95% were married or living with a partner and 66% had a degree or higher professional qualification. 86% lived in a house or bungalow and 13% in a flat. There were no significant differences in background characteristics between BL and DL mothers. Compared to women included in this analysis, those who did not provide details of their delivery experience and infant feeding were significantly younger, less likely to self-identify as white, less likely to be married or living with a partner, less likely to have a degree or higher qualification and more likely to have a male infant (data not shown).

**Table 1.** Background characteristics of women who completed the survey in week 1, according to whether they delivered before or during the lockdown (mean(SD) or n(%))

|  | **Baby born before lockdown**<br>**n=1049** | **Baby born during lockdown**<br>**n=316** |
| --- | --- | --- |
| Maternal age (y) | 31.7 (4.6) | 31.4 (4.9) |
| Infant age (mo) | 5.9 (2.7) | 1.27 (0.76)** |
| Infant gestation (w) | 39.3 (1.8) | 39.4 (1.7) |
| Male infant | 535 (51) | 156 (49) |
| Other children in household | 574 (55) | 169 (54) |
| *Maternal education* |  |  |
| ≤5 GCSE A-C grade | 46 (4) | 13 (4) |
| ≥5 GCSE A-C grade | 85 (8) | 22 (7) |
| A levels//equivalent | 227 (22) | 60 (19) |
| Bachelors degree | 400 (38) | 134 (42) |
| Master's degree | 144 (14) | 44 (14) |
| PhD/professional qualification | 140 (13) | 41 (13) |
| *Current accommodation* |  |  |
| Flat/apartment | 139 (13) | 37 (12) |
| House/bungalow | 899 (86) | 273 (86) |
| *Household income* |  |  |
| < £20K | 74 (7) | 17 (5) |
| < £30K | 111 (11) | 32 (10) |
| < £45K | 187 (18) | 53 (17) |
| < £75K | 334 (32) | 107 (34) |
| < £100K | 154 (15) | 50 (16) |
| >£100K | 143 (14) | 40 (13) |
| Prefer not to say | 40 (4) | 16 (5) |

y: years; mo: months; w weeks
\*\*p<0.001

**Figure 1.**
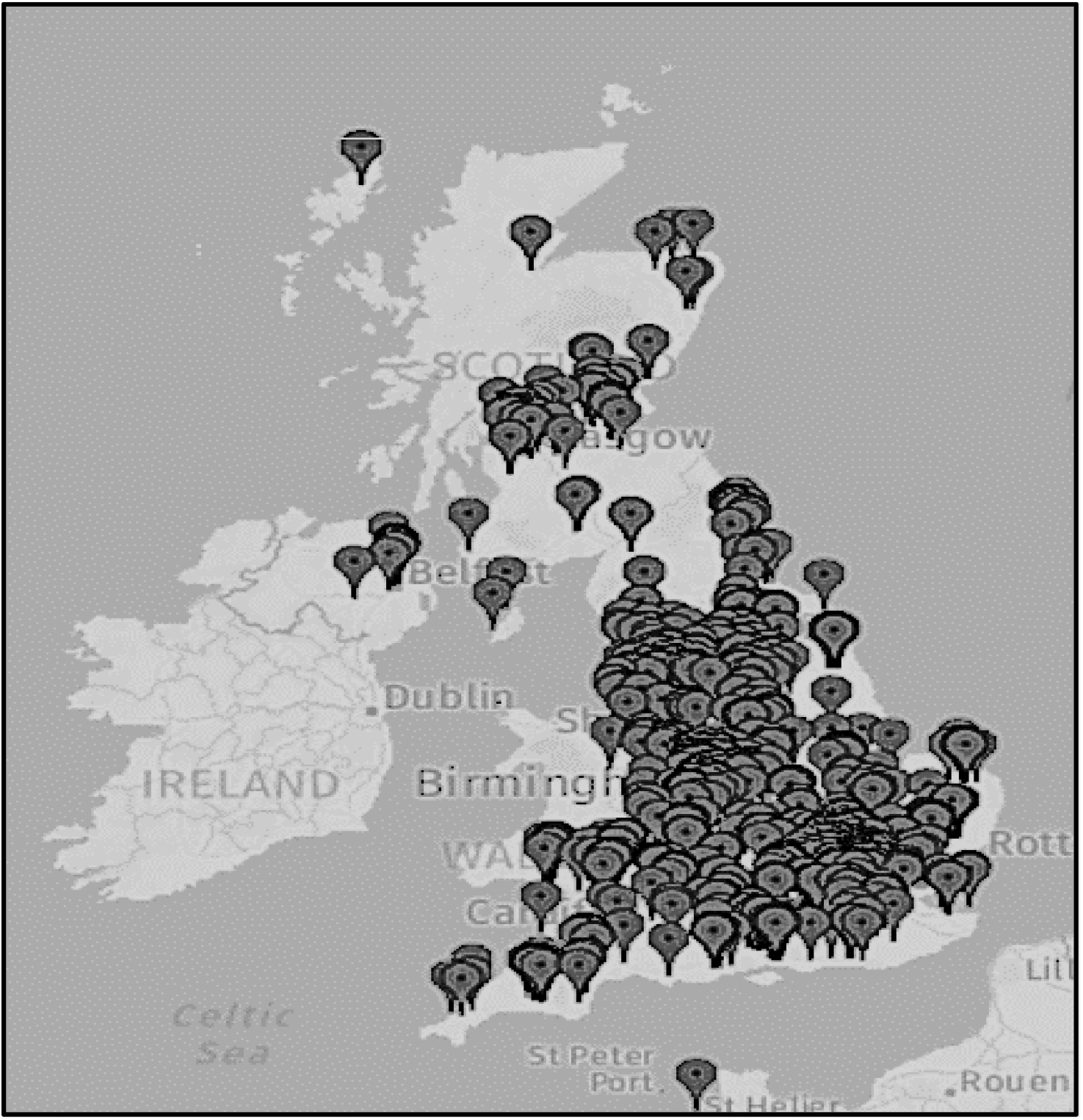
Geographical location of survey participants who completed the survey in week 1 (by first 3 characters of postcode)

### Birth experiences

The mode of delivery did not differ between BL and DL groups. BL women spent significantly more nights in hospital (p<0.001) reflecting fewer long stays in DL mothers (Figure 2). A high proportion of those in both groups practiced skin-to-skin contact shortly after delivery, with no difference in timing or duration between groups. Most women intended to breast-feed and initiated breastfeeding within the first hour after delivery. Seventy-six versus 72% of BL and DL women reported receiving enough help and support with feeding whilst in hospital, and 71 versus 67% had help from a health professional in positioning the infant during breastfeeding (Table 2).

**Table 2.** Birth and early feeding experiences of women who completed the survey in week 1, according to whether they delivered before or during the lockdown (n (%))

|  | **Baby born before**<br>**lockdown n=1049** | **Baby born during**<br>**lockdown n=316** |
| --- | --- | --- |
| *Mode of delivery* |  |  |
| Vaginal | 473 (45) | 142 (45) |
| Vaginal induced | 258 (25) | 96 (30) |
| Planned caesarean | 122 (12) | 34 (11) |
| Unplanned/Emergency caesarean | 196 (19) | 44 (14) |
| *Birth location* |  |  |
| Home | 51 (5) | 17 (3) |
| NHS Hospital | 987 (94) | 298 (94) |
| *Skin-to-skin contact after delivery* | 914 (87) | 281 (89) |
| *Initiation of skin-to-skin contact* |  |  |
| Immediate | 674 (74) | 210 (75) |
| <30mins | 182 (20) | 58 (21) |
| >30mins | 40 (4) | 6 (2) |
| >1hr | 17 (2) | 7 (3) |
| *Duration of skin-to-skin contact* |  |  |
| <30 mins | 234 (26) | 70 (25) |
| 30-60 mins | 299 (33) | 102 (36) |
| >1hr | 378 (41) | 109 (39) |
| *Feeding intention before delivery* |  |  |
| Breastfeeding | 897 (86) | 281 (89) |
| Formula feeding | 67 (6) | 7 (2) |
| Mixed feeding | 73 (7) | 25 (8) |
| No plans | 9 (1) | 2 (1) |
| *Timing of initiation<sup>+</sup>* |  |  |
| Did not breastfeed | 9 (1) | 4 (1) |
| <30 mins | 411 (47) | 119 (43) |
| 30-60 mins | 309 (35) | 108 (39) |
| 1-2hrs | 154 (17) | 49 (18) |
| *Received enough help with breastfeeding in hospital<sup>+</sup>* |  |  |
| Yes | 754 (76) | 212 (72) |
| No | 237 (24) | 85 (28) |
| *Received enough help with positioning in hospital<sup>+</sup>* |  |  |
| Yes | 697 (71) | 198 (67) |
| No | 283 (29) | 99 (33) |

+ of those planning to breastfeed

**Figure 2.**
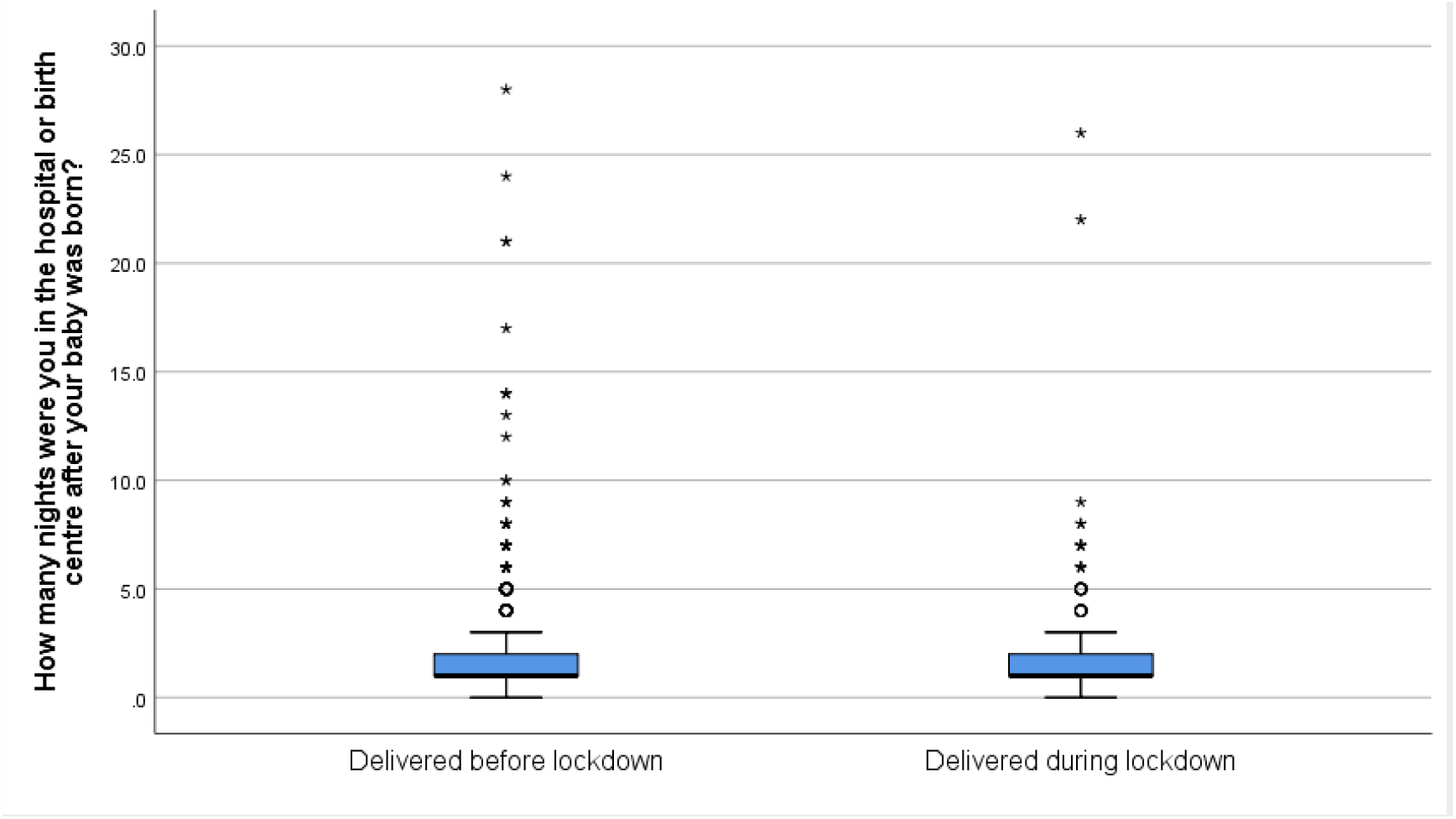
Number of nights spent in hospital for women who delivered before versus during lockdown and completed the survey in week 1.

Among DL women, 39% reported that their birth plan had changed due to the pandemic. The main changes reported as free-text (more than one could be given) were: (1) Having to give birth in hospital rather than in a low-risk/ midwife-led unit (34 women); (2) Only being allowed one birth partner (24 women); (3) Limited support from their birth partner, who could only be present during active labour but not during induction, early labour or after delivery (40 women); (4) No partner being present (11 women; various reasons given for this including problems with childcare); (5) Water birth not being available (11 women); (6) Not being allowed a home birth (9 women).

### Infant feeding

Infant feeding practices at the time of survey completion are shown in Table 3. As expected, reflecting the younger age of the infants of DL mothers, a significantly higher proportion were exclusively breast-fed or fed a combination of breast milk and infant formula (59% versus 39%), whereas a higher proportion of older infants born before lockdown were also consuming solid foods.

**Table 3.** Infant feeding and postnatal support experiences of women who completed the survey in week 1, according to whether they delivered before or during the lockdown (n (%))

|  | Baby born before lockdown<br>n=1049 | Baby born during lockdown<br>n=316 |
| --- | --- | --- |
| *Current infant feeding* |  |  |
| Exclusive breastfeeding | 323 (31) | 214 (40)** |
| Formula feeding | 130 (12) | 43 (14) |
| Mixed milk feeding | 80 (8) | 59 (19) |
| Breastfeeding + solids | 335 (32) | 0 |
| Formula feeding plus solid | 138 (13) | 0 |
| Mixed feeding plus solids | 43 (4) | 0 |
| *Who supports infant feeding?+* |  |  |
| Partner | 623 (59) | 198 (63) |
| Parent | 225 (21) | 65 (21) |
| In-laws | 53 (5) | 20 (6) |
| Health professional | 518 (49) | 169 (54) |
| Friend/relative | 394 (38) | 110 (35) |
| BF support group | 345 (33) | 97 (31) |
| Online support | 510 (49) | 136 (43) |
| Other | 46 (4) | 14 (4) |
| No support | 109 (10) | 36 (11) |
| *Who has the greatest influence on infant feeding?+* |  |  |
| Partner | 399 (38) | 118 (37) |
| Parent | 94 (9) | 30 (10) |
| In-law | 12 (1) | 6 (2) |
| Health professional | 229 (22) | 78 (25) |
| Friend/relative | 217 (21) | 55 (17) |
| Support group | 197 (19) | 59 (19) |
| Online support | 228 (22) | 49 (16)* |
| Other | 30 (3) | 13 (4) |
| *Contact with a health professional (per week)* |  |  |
| Never | 699 (67) | 104 (33)** |
| 1-3 times | 312 (30) | 186 (59) |
| 4-5 times | 14 (1) | 18 (6) |
| Daily or more | 1 (<1) | 4 (1) |

\*p<0.05, \*\*p<0.005

**Table 3. Continuation**
|  | **Baby born before lockdown**<br>**n=1049** | **Baby born during lockdown**<br>**n=316** |
| --- | --- | --- |
| *Contact with a Mother&Baby or breastfeeding support group (per week)* |  |  |
| Never | 780 (74) | 211 (67)* |
| 1-3 times | 165 (16) | 74 (23) |
| 4-5 times | 40 (4) | 13 (4) |
| Daily or more | 43 (4) | 15 (5) |
| *Contact with a mental health professional (per week)* |  |  |
| Never | 963 (92) | 288 (91) |
| 1-3 times | 58 (6) | 23 (7) |
| 4-5 times | 7 (1) | 3 (1) |
| Daily or more | 1 (<1) | 0 |
| *Enough support with own health?* |  |  |
| Yes | 624 (60) | 175 (55) |
| No | 423 (40) | 140 (44) |

\*p<0.05, \*\*p<0.005

Thirteen % of mothers in both groups reported having changed their mode of infant feeding in response to the lockdown. Of the mothers who were breastfeeding, 60% reported no change in feed frequency, whilst 30% reported an increase and 10% a decrease. 68% reported no change in the duration of feeds, whilst 17% reported an increase and 15% a decrease. Four % of women reported they had stopped breastfeeding. Of the mothers who were formula feeding, 66% reported no change, 18% an increase, 13% a decrease and 11% that they had stopped formula feeding during lockdown. Most women reported no change to their plans for introducing solid foods as a result of the lockdown (89%), although 8% had introduced solid foods earlier than planned and 3% later.

DL mothers had the opportunity to provide free-text responses about how their feeding plans had changed. The most frequent responses related to a lack of breastfeeding support (n=21), especially face-to-face help with practical problems such as latching, resulting in the mother expressing milk, introducing formula or stopping breastfeeding. Six women reported that they had breastfeeding problems because their infant had a tongue-tie which could not be dealt with surgically due to the pandemic. Seven women reported a change to both their birth plan and feeding plans.

The main reported sources of infant feeding support in both groups of women (‘where do you get support with infant feeding’) were the partner (60%), health professional (50%) or an online support group (such as Facebook, 47%), followed by friends and family (37%), and infant feeding support groups such as NCT, La Leche league (32%). There were no significant differences between groups. In free-text responses under the ‘other’ category, 28 women mentioned support from a lactation consultant or counsellor. In responses to the question ‘who is most helpful or influential with infant feeding’, the highest proportion of women reported this was their partner (38%), with lower but similar results for health professionals (20%), friends and family (19%), support groups (19%) and online groups (20%). Significantly fewer women from the DL group reported that online groups were a main influence on infant feeding compared to those from the BL group (16% v 22%, p=0.02).

Fifty-nine % of DL women reported that they had received professional help with breastfeeding in the first few days after delivery, whilst 40% received support from family and friends. In response to the question ‘ did you feel you got or are getting enough support and help with feeding your baby’, overall, 45% of DL mothers felt they had not received enough support and help in feeding their infant from delivery to completing the survey.

Among BL women, 57% reported that their infant feeding support had decreased since lockdown, with 20% reporting no change, 1% an increase and 21% that this did not apply to them. Childcare support was also reported to have decreased by 69% of participants.

Mothers were able to provide free-text information on how COVID-19 had affected them. In response to this, 146 women (11%) indicated that lockdown had adversely affected their mental health, citing anxiety, depression, isolation and loneliness. 119 women (9%) mentioned consequences of not being able to see their family, highlighting the lack of practical support but also distress that family members had missed seeing the new baby; 62 (5%) highlighted the lack of social support from their friends, and missing attending mother and baby groups and activities; while 26 (2%) highlighted the lack of face to face GP and health visitor visits. Several women mentioned that their experience of late pregnancy, birth and early motherhood had been completely different from what they had expected and that they and their extended family were missing out on experiences that could never be regained. Two women also highlighted positive aspects of increased bonding with their other children.

Contact with a health professional during lockdown was significantly more frequent in DL mothers, who had younger infants than those in the BL group. Similarly, these women were more likely to have had contact with a mother and baby or breastfeeding support group (33% versus 26%, p=0.018). Of the 350 women who reported these contacts, 9 episodes were reported to have been ‘in person’ with the rest online or by phone. 92 women (8% BL and 9% DL) reported an appointment with a mental health professional; four in person, 11 both in person and remotely, and the rest just remotely. The proportion of women who reported that they received enough support with their own health was 60% versus 55% for BL and DL groups.

## Discussion

The COVID-19 pandemic and lockdown has affected every facet of day-to-day life. Our preliminary analysis of data from the New Mum Study, including participants from most parts of the UK, indicates that for women and their infants, this ‘new reality’ affects their experiences during delivery and after birth.

### Birth experience and early feeding

Our data suggest that, despite difficulties imposed by the pandemic, hospital facilities are continuing to implement measures such as promoting early mother-baby contact and initiation of breastfeeding, thus following guidelines that encourage these practices during the pandemic ^2-4^. Eighty-seven % of BL mothers and 89% of DL mothers practised skin-to-skin contact, and almost all did so within the first hour after birth. Similarly, 86% of BL mothers and 89% of DL mothers intended to breastfeed, and 82% initiated breastfeeding within the first hour after delivery. These figures are higher than reported in the 2010 National Infant Feeding Survey where 81% reported skin-to-skin contact and 74% initiated breastfeeding in the first hour^5^. It is possible that these practices have improved during the intervening years although it is perhaps more likely to reflect the characteristics of the survey participants who are not currently representative of the general UK population. Nevertheless, the maintenance of these important hospital practices since the start of the COVID-19 pandemic and UK lockdown is a positive step in supporting breastfeeding. Keeping mother and infant together and avoiding disruption of breastfeeding due to COVID-19 infection concerns is important in minimising psychological stress and detrimental effects on feeding and bonding ^6^.

Despite these positive practices, about a third of the women who planned to breast-feed reported that they did not receive help with positioning the infant and a quarter perceived that they did not get enough support with feeding in the hospital. This could be due to the increased burden on healthcare systems and pressure placed on healthcare professionals to discharge mothers sooner to minimise infection risks ^6-7^, which might result in less opportunity to support mothers with infant feeding. The shorter length of hospital stays after births during the lockdown compared to those before the lockdown might provide further evidence of this. However, since there was no significant difference in the support received by mothers who delivered before or during the lockdown, they may not reflect specific effects of the lockdown. Furthermore, the figures for help with positioning are similar to those reported in the 2010 Infant Feeding Survey.

Having a birth plan encourages women to communicate their expectations related to labour and delivery and may give them a sense of personal achievement and control through involvement in decision-making ^8-9^. The current situation is inevitably affecting women’s decisions about their birth plans and this lack of control could negatively affect their birth experience ^9^. Our analysis showed that 39% of women who delivered since lockdown reported that their birth plans had changed due to COVID-19. One of the most frequent examples was having to give birth in a hospital rather than in a midwife-led unit, which could increase their risk of infection ^10^. Practices in the United Kingdom during the pandemic vary, with some services maintaining and expanding their midwife-led unit services while others are moving towards centralization ^10^.

Another commonly reported change to the birth plan was being limited to one birth partner, and the partner only being present during active labour. UK practices are less strict than those in several European countries that introduced rules prohibiting birth supporters during labour and/or in the postpartum period ^10^. Minimising the number of caregivers the infant and mother are exposed to is important to reduce infection risk ^11^. However, continuous support to women during childbirth, which can be provided by midwives, nurses, doulas, partners, family or friends, may lead to improved outcomes for women and infants, including increased spontaneous vaginal delivery, shorter labour duration and decreased use of analgesia, higher five-minute Apgar score and a reduction in negative feelings about the childbirth experience ^12^. The absence of the expected emotional and practical support from birth partners resulting from measures to protect mothers and infants from infection could thus be affecting other maternal and childbirth outcomes.

### Infant feeding practices

Overall, breastfeeding rates in the survey population appear relatively high, but direct comparisons with UK data are difficult given the wide age-range of infants. 13% of women reported changes to infant feeding as a result of the lockdown. Of women who were breastfeeding, 30% reported an increase in the frequency and 17% an increase in the duration of feeds, which could reflect more time spent at home, experiencing more frequent support from the partner and/or being able to invest more time in childcare. Conversely, 10% of women reported a decrease in breastfeeding frequency and 15% a decrease in feed duration. Thus the effects of the pandemic on infant feeding may vary depending on the context, access to support and special circumstances experienced by individual mothers. For example, a report by the Institute of Fiscal Studies ^13^ highlighted that COVID measures have affected men and women’s workloads and responsibilities in different ways and suggested a greater investment by mothers in domestic responsibilities and childcare. This issue will be further investigated in future analyses of survey data.

### Infant feeding support

Support from relatives, friends and other social networks (e.g. health care professionals and online groups) can help to promote the skills needed to overcome adversity and other mental health challenges ^14-15^ that could further complicate maternal decisions about infant care. Of mothers who delivered during the lockdown, 45% felt they were not getting enough support with feeding (beyond hospital assistance), whilst 57% of those who delivered before lockdown had experienced a decrease in infant feeding support during this period. This is of concern given evidence that the quality of breastfeeding support is important for maternal mental health. In a prospective study of Canadian women with breastfeeding difficulties, those who did not report a negative breastfeeding support experience were at decreased risk of postpartum depression ^16^.

Women from both groups reported that their partner was the greatest source of support and also had the greatest influence on infant feeding. Potentially, a partner who seeks more active participation in childcare during this exceptional period could represent a valuable source of support for women, especially for those who are experiencing limited access to other family members or friends who might provide this support under normal circumstances.

Reliance on support from online groups was significantly lower for women who delivered during lockdown, possibly because their infants were younger and they needed more practical support for the establishment of breastfeeding or other aspects of care. Women from this group also reported more frequent contact with health professionals.

Finally, 11% of mothers reported an impact of the lockdown measures on their mental health, mentioning anxiety, depression, loneliness and isolation. A similar proportion reported they had consulted a mental health professional. This is consistent with the estimated 10-20% of women in England and Wales who develop a mental health problem during the perinatal period, although this includes pregnancy and the first year post-partum, so the figure is not directly comparable with that from our survey^17^. Lack of face-to-face contact with relatives was also mentioned, both in the context of reduced practical support but also distress at not being able to share early motherhood experiences and introduce the new baby. Responses expressed a sense of feeling cheated of the anticipated experience of birth and motherhood, something that would not ever be regained. Emotional distress in women may have short-term consequences, with inhibition of the let-down reflex leading to disruption of milk flow and reduced milk volume^18,19^. The longer-term significance of these findings for mother-infant interactions and subsequent outcomes is unclear, but merits further investigation.

## Strengths and limitations

By using an anonymous online survey we have been able to collect information rapidly from a large group of women from all regions of the UK. However, a significant limitation highlighted by this analysis is that the participants are not currently representative of all new mothers in the UK. Compared to national data, our participants have higher educational attainment compared to the 2011 Census figures for England & Wales showing 37% of mothers have no qualifications or less than 5 GCSEs or equivalent^20^. They have a higher income when compared to the Family Resources Survey 2018/19 which shows roughly 40% of families with children in the UK have a gross income above £1000/week or approximately over £52000 a year^21^. Fewer of our participants come from a BAME background compared to 2011 Census figures which indicate that the prevalence of adult women of Asian, Black, Mixed, White and Other ethnicities in England & Wales are 6.8%, 3.0%, 1.4%, 88% and 0.8% respectively^20^. Finally, a higher proportion of participants are married or cohabitating mothers compared to 201<u>9</u> ONS data which shows that in the UK roughly 67% of families with dependent children are married or in a civil partnership, 17% are cohabitating, 2% are lone father and 15% are lone mothers^22^.

There is evidence that COVID-19 and resulting lockdown measures disproportionately affect BAME and disadvantaged groups^23^. Women in BAME groups are also at higher risk of becoming significantly unwell with COVID-19 in pregnancy and requiring hospital admission, although this study did not look at other socio-economic differences^24^. COVID-19 and lockdown measures may also exacerbate existing food inequalities for families with children^25^. While many explanations have been advanced for the observed COVID-19 ethnic disparities, it is likely that pre-existing social and health inequalities and differences in the usage of and need for health services are an important factor ^26,27^. It is vital that the experiences of these groups are represented in the survey, and we are taking advice and joining efforts with other groups to reach these groups and increase the representativeness of our survey sample.

### Immediate implications and further analyses

Our findings highlight the impact of the current pandemic and lockdown measures on the birth experiences, infant feeding and support experienced by mothers. Some practical steps suggested by the findings are improving infant feeding support, especially ‘face-to-face’ support for practical issues such as latching and recognising and supporting mothers who are struggling with mental health challenges or indeed other aspects of their health. Interestingly, a recent qualitative study in 14 women^28^, 11 with young infants, reported similar issues regarding feelings of loss and the lack of face-to-face support. It highlighted the ‘digital pivot’ that is taking place with perinatal support organisations moving online. However, the effectiveness of online versus face-to-face contact is currently uncertain, and it may not be accessible to all women. Ultimately, it is likely that some of the challenges being reported by mothers will be alleviated by the relaxation of lockdown measures so that they can resume contact with their extended families and friendship groups. However, it is also important to consider whether the altered birth experiences and challenges experienced by these women will have later consequences for them and/or their infants.

Future analyses of the survey data will explore whether experiences change as restriction measures lift, ideally in a larger and more diverse sample. Analyses will also include more data about participants’ living conditions as well as the effects of the lockdown on everyday life, finances and maternal mood and how these factors are associated with infant feeding decisions and support.

## Data Availability

The full survey dataset will be made available when the survey is closed, via the study webpage https://www.ucl.ac.uk/child-health/research/population-policy-and-practice-research-and-teaching-department/cenb-clinical-11.

## Conflict of interest statement

The authors declare no conflict of interest with respect to the COVID-19 New Mum Study or this manuscript. MF receives an unrestricted donation for research on infant nutrition from Philips. The remaining authors declare no other conflicts.

## Notes

### Funding Statement

No funding was received for the survey. All research at Great Ormond Street Hospital NHS Foundation Trust and UCL Great Ormond Street Institute of Child Health is made possible by the NIHR Great Ormond Street Hospital Biomedical Research Centre. The views expressed are those of the author(s) and not necessarily those of the NHS, the NIHR or the Department of Health.

### Author Declarations

Ethical approval was obtained from the UCL Research Ethics committee (0326/017).

